# Patterns of COVID-19 related excess mortality in the municipalities of Northern Italy

**DOI:** 10.1101/2020.05.11.20097964

**Authors:** Dino Gibertoni, Kadjo Yves Cedric Adja, Davide Golinelli, Chiara Reno, Luca Regazzi, Maria Pia Fantini

## Abstract

The Coronavirus Disease 2019 (COVID-19) spatial distribution in Italy is inhomogeneous, because of its ways of spreading from the initial hotspots. The impact of COVID-19 on mortality has been described at the regional level, while less is known about mortality in demographic subgroups within municipalities.

We aimed to describe the excess mortality (EM) due to COVID-19 in the three most affected Italian regions, by estimating EM in subgroups defined by gender and age classes within each municipality from February 23 to March 31, 2020.

EM varied widely among municipalities even within the same region; it was similar between genders for the ≥75 age group, while in the other age groups it was higher in males. Thus, nearby municipalities may show a different mortality burden despite being under common regional health policies, possibly as a result of policies adopted both at the regional and at the municipality level.

## 1. Introduction

As of May 10, 2020, 4,051,431 confirmed cases and 279,734 deaths of Coronavirus Disease 2019 (COVID-19), a respiratory infectious disease caused by the Severe Acute Respiratory Syndrome - Coronavirus 2 (SARS-CoV-2), were reported worldwide, affecting 187 countries (Center for Systems Science and Engineering (CSSE) at Johns Hopkins University (JHU), 2020). Among Western countries, Italy was the first affected. The first case of local transmission of COVID-19 was confirmed in a 38-year-old man in the municipality of Codogno (Lombardy region) on February 20, 2020 (Zangrillo et al., 2020), while on February 21, a resident of Vo’, a small town near Padua (Veneto region), died of pneumonia due to SARS-CoV-2 infection (Crisanti and Cassone, 2020; Lavezzo et al., 2020). On February 23, the Italian government created two red zones confining ten towns including Codogno and the town of Vo’, quarantining 50,000 people.

Despite these containment measures, SARS-CoV-2 rapidly spread not only in the Lombardy and Veneto region, but also in some areas of the bordering Emilia Romagna region. It became clear that, without appropriate measures, SARS-CoV-2 would have spread dangerously all over Italy. This prospect convinced the government to lockdown the whole country on March 10, 2020, when confirmed cases were 10,149 and 631 deaths (Italian Civil Protection Department, 2020). At first, commercial activities were closed, but some productive ones were allowed to stay open. On March 22, the government closed all productive activities, except for those necessary to produce essential goods. As of May 10, 2020, official cases in Italy are 219.070 (Italian Civil Protection Department, 2020), while deaths from COVID-19 reached 30.560. The COVID-19 had an extremely varied distribution in Italy, because of its ways of spreading and of geographical differences in population density, distribution of healthcare facilities, presence of care and nursing homes and locally defined strategies to contain/mitigate its spreading. As a matter of fact, in Italy, regions must provide the so called “Essential Levels of Care” (i.e. the core benefit package and standard of health services), set by the central government, through the Regional Health Service (RHS), but they are responsible and autonomous for the local organization and delivery of health care (Lenzi et al., 2013). In each region, the municipalities followed the general indications given by the Regional Government, but with different scaling up strategies at the local level.

Deaths from COVID-19 as reported by official data may be underestimated because of various factors, among which the classification of cause of death when the deceased suffered from other pre-existing conditions and the absence of post-mortem testing among those who have died before they could be swabbed. For this reason, the most reliable method to estimate COVID-19 related mortality to date is to calculate the excess all-cause mortality occurred during the COVID-19 outbreak compared to all-cause mortality in the same period in the previous years. The Italian National Institute of Statistics (ISTAT), called on by researchers and local administrators, promptly released and regularly updated the data of confirmed deaths (Italian Institute for National Statistics (ISTAT), 2020b). The first studies (Colombo and Impicciatore, 2020; Mancino, 2020) performed on these data reported higher excess mortality mainly in the municipalities closer to the SARS-CoV-2 outbreak epicenters, such as those in Lombardy. Most of these studies (Bucci et al., 2020; Buonanno et al., 2020; Modi et al., 2020) aimed to infer excess mortality at the regional or national level, but all of them had to acknowledge the caveats implied by relying on incomplete data to draw such predictions. Other studies, utilizing data provided by the heat waves surveillance systems collected only in a limited number of municipalities (Davoli et al., 2020), provided conflicting results, without showing a particular excess of mortality.

Importantly, excess mortality is not evenly distributed over the national territory and at the regional and municipality level (Davoli et al., 2020; Italian Institute for National Statistics (ISTAT), 2020b; Italian Institute for National Statistics (ISTAT) and Italian Superior Institute of Health, 2020; Mancino, 2020; ScienzaInRete, 2020). In fact, some municipalities seem to be unaffected by the epidemic, with a number of deaths equal to or even lower than that related to the same period of previous years, while in others the number of deaths was estimated between 6 to 10 times higher.

Therefore, the aim of our study is to describe the spatial and demographic distribution of excess mortality due to COVID-19 in the three most affected Italian regions, by estimating excess mortality in subgroups defined by gender and age classes within each municipality with available data.

## 2. Methods

We conducted a longitudinal study using the data provided by the Italian Institute of Statistics (ISTAT) on May 4, 2020, which include the number of deaths available for 6,866 Italian municipalities (87.0% out of the 7,904 municipalities) (Italian Institute for National Statistics (ISTAT), 2020c) divided by gender and age classes in the period from January 1 to March 31 of the years 2015-2020. The municipalities with available data were 1,439/1,506 (95.8%) in Lombardy, 490/563 (87.0%) in Veneto and 295/328 (89.9%) in Emilia-Romagna.

Excess mortality (EM) was estimated for each municipality in six subgroups defined by gender and three age classes consistent with data provided by ISTAT (0-64, 65-74 and ≥75 years) in the time period from February 23, 2020 to March 31, 2020, corresponding to the EPidemic Period (EPP). In each subgroup a simple linear regression was conducted on the observed deaths from 2015 to 2019; the constant and slope obtained were used to estimate by extrapolation the expected number of deaths in 2020. The ratio of observed to expected death is the relative mortality (RM) experienced by each subgroup and the corresponding excess mortality is obtained by subtracting 1 from the RM and multiplying this value by 100. To avoid obtaining a negative expected number of deaths, which might happen in the smallest municipalities, we set to 1 each prediction ≤0. Moreover, taking into account that 2020 is a leap year we corrected estimates by dividing for a coefficient of 1.022.

To summarize the impact of COVID-19, we reported for each age and gender subgroup how many municipalities showed an excess mortality (separating those under +50%, which could include also not anomalous increases of mortality, from those over +50%) and how many showed a lower mortality in the EPP.

Lastly, relative mortality ratios were applied to choropleth maps (Pisati, 2007) of the three Italian regions considered, in order to visually identify the areas that suffered the greatest burden of COVID-19 mortality during the epidemic and the municipalities where a reduction in mortality was observed. We drew side by side regional maps of two age groups (0-64, ≥75 years) to visually check the RM differences.

All analyses were carried out using Stata v.15.1 (StataCorp; College Station, TX, USA).

## 3. Results

Relative mortality during the COVID-19 outbreak exhibited a very composite pattern among the three examined regions, and between age classes and genders (Table 1). Generally, Lombardy experienced the highest burden of mortality, and Veneto the lowest. In the three regions, females displayed an excess mortality only among those aged ≥75 years. Males in age groups 65-74 and ≥75 had excess mortality in the three regions, except for the age group 65-74 in Veneto. People aged ≥75 years had the highest excess mortality. In Lombardy, this excess was +170% for males and +146% for females, in Emilia-Romagna +123% for males and +104% for females; in Veneto +35% for males and +37% for females. While among people aged ≥75, EM was quite similar between males and females, in the other two age classes it was higher in males. For instance, in the 65-74 age group the EM of males was almost twofold compared to females in Lombardy and Emilia Romagna.

**Table 1.** Relative mortality in municipalities (age and gender subgroups) during the COVID-19 epidemic. Median, mean and maximum values are reported for each subgroup.

| LOMBARDIA (n=1439) | EPP<br>(23.02.2020 to 31.03.2020) |  |  |
| --- | --- | --- | --- |
|  | median | mean | maximum |
| 0-64y Females | 0 | 0.43 | 7 |
| 0-64y Males | 0.29 | 0.87 | 10 |
| 65-74y Females | 0 | 0.71 | 9 |
| 65-74y Males | 1 | 1.57 | 22 |
| $\geq 75$y Females | 1.5 | 2.46 | 36 |
| $\geq 75$y Males | 2 | 2.70 | 47 |

| VENETO (n=490) | EPP<br>(23.02.2020 to 31.03.2020) |  |  |
| --- | --- | --- | --- |
|  | median | mean | maximum |
| 0-64y Females | 0 | 0.36 | 3 |
| 0-64y Males | 0 | 0.52 | 5 |
| 65-74y Females | 0 | 0.39 | 5 |
| 65-74y Males | 0 | 0.64 | 6 |
| $\geq 75$y Females | 1 | 1.37 | 11 |
| $\geq 75$y Males | 1 | 1.35 | 13 |

| EMILIA-ROMAGNA (n=295) | EPP<br>(23.02.2020 to 31.03.2020) |  |  |
| --- | --- | --- | --- |
|  | median | mean | maximum |
| 0-64y Females | 0 | 0.47 | 5 |
| 0-64y Males | 1 | 1.02 | 26 |
| 65-74y Females | 0.5 | 0.82 | 6 |
| 65-74y Males | 1 | 1.38 | 8 |
| $\geq 75$y Females | 1.43 | 2.04 | 14 |
| $\geq 75$y Males | 1.53 | 2.23 | 14 |

### 3.1. Municipality level

In Lombardy, males aged ≥75 experienced an EM in 60.3% of municipalities, compared to the previous five years, while in the remaining 39.7% no EM was observed (Table 2). In Emilia-Romagna the proportion of municipalities with EM was similar to that of Lombardy (64.4% of municipalities, males ≥75 years) while in Veneto it was lower (40.6%). These figures were similar for females aged ≥75. As illustrated in Table 2, in most of the municipalities no EM was found for males and females in the age groups 0-64 and 65-74.

**Table 2.**
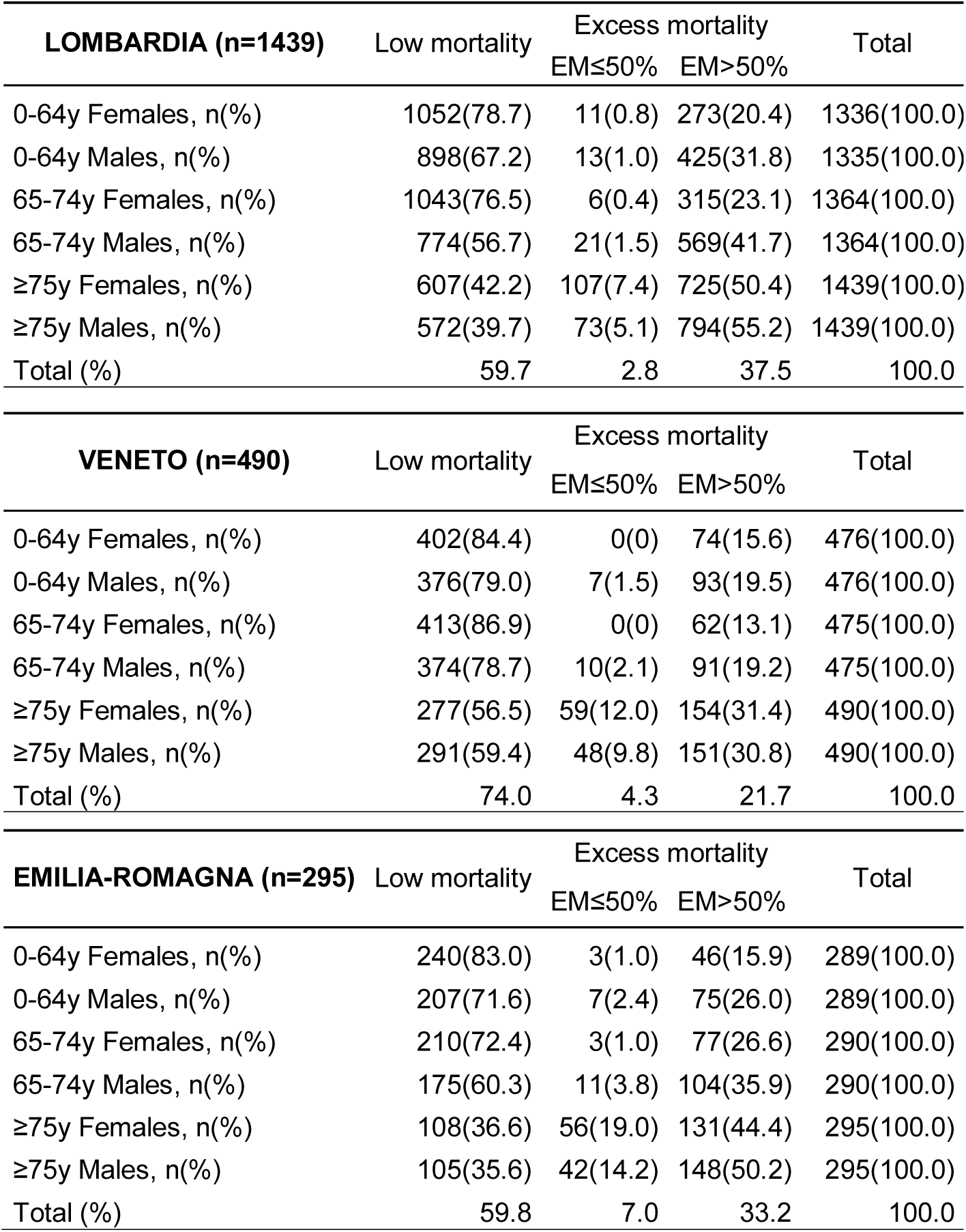
Cross-classification of municipalities (age and gender subgroups) with low or excess mortality during the COVID-19 outbreak. The number (and percentage) of municipalities for each subgroup is reported.

Concerning individual municipalities (Supplementary materials), the highest EM was estimated in Brembate di Sopra (10 km from Bergamo, in Lombardy) in males aged ≥75 (+4,600%), with 47 deaths occurred against 1 expected.

In the major towns, Parma (Emilia-Romagna) displayed a +633% EM in males aged 65-74; Bergamo (Lombardy) +551% EM in males aged ≥75, +409% EM in males aged 65-74 and +368% EM in females aged ≥75; Piacenza (Emilia-Romagna) had +491% EM in males ≥75. The regional capitals Venezia (Veneto) and Bologna (Emilia-Romagna) experienced EM rates generally below +50%. On the contrary, Milano (Lombardy) had an EM of +166% in males aged 65-74 and an EM between +56% and +76% in the other groups except for females aged 0-64 with +4%.

Examination of the Lombardy map for men aged ≥75 (Figure 1) confirmed that the municipalities of Bergamo, Brescia, Cremona and Lodi provinces, corresponding to the central and eastern part of the region, experienced the highest EM. The “green” municipalities with a mortality reduction are mainly located in the peripheral areas of the region which are more distant and less connected to the epidemic epicenters. In men aged 0-64 this pattern is less visible, because most municipalities did not experience an EM for this age group. Differently from Lombardy, a geographical pattern of epidemic spread for males aged ≥75 is not clearly visible in Veneto (Figure 2), as municipalities with EM seem to be scattered in different areas, intertwined with “green zones”. Moreover, in men aged 0-64 very few municipalities displayed an EM. The western Verona province, bordering Lombardy, was not the most hit by mortality within the region. From these maps we can argue that Veneto successfully isolated itself from the potential epidemic flow coming from the nearby region.

**Figure 1.**
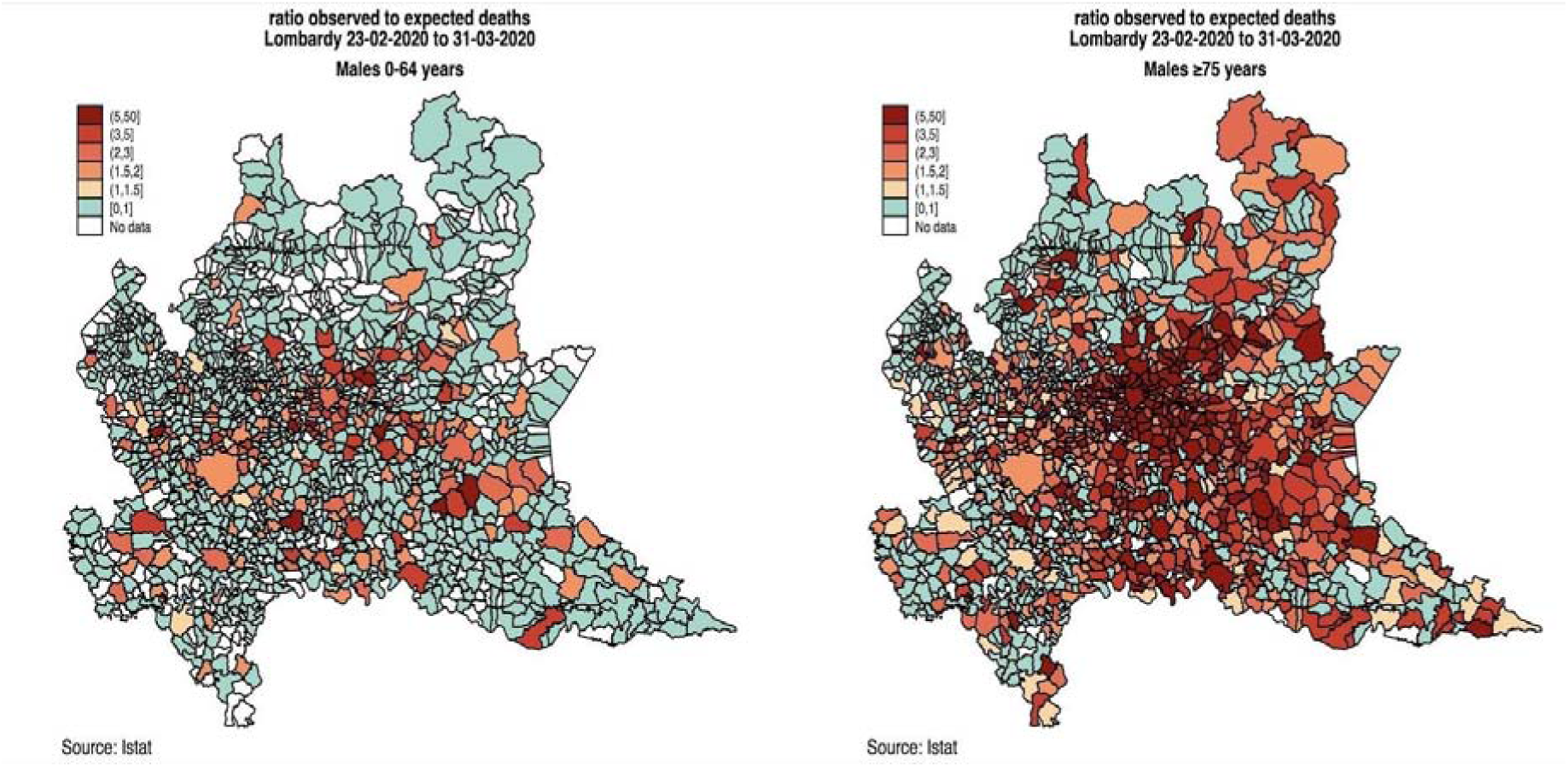
Relative mortality in males aged 0-64 (left) and ≥75 years (right) in Lombardy region. Municipalities are depicted with different colors according to the magnitude of their excess mortality. Green refers to municipalities with a relative mortality ≤1, that is when observed deaths are lower than or equal to the expected deaths. When the relative mortality is >1, increasing saturation of red is used, to reflect increasing values of relative mortality. Municipalities depicted in white are those for which no data was released by ISTAT.

**Figure 2.**
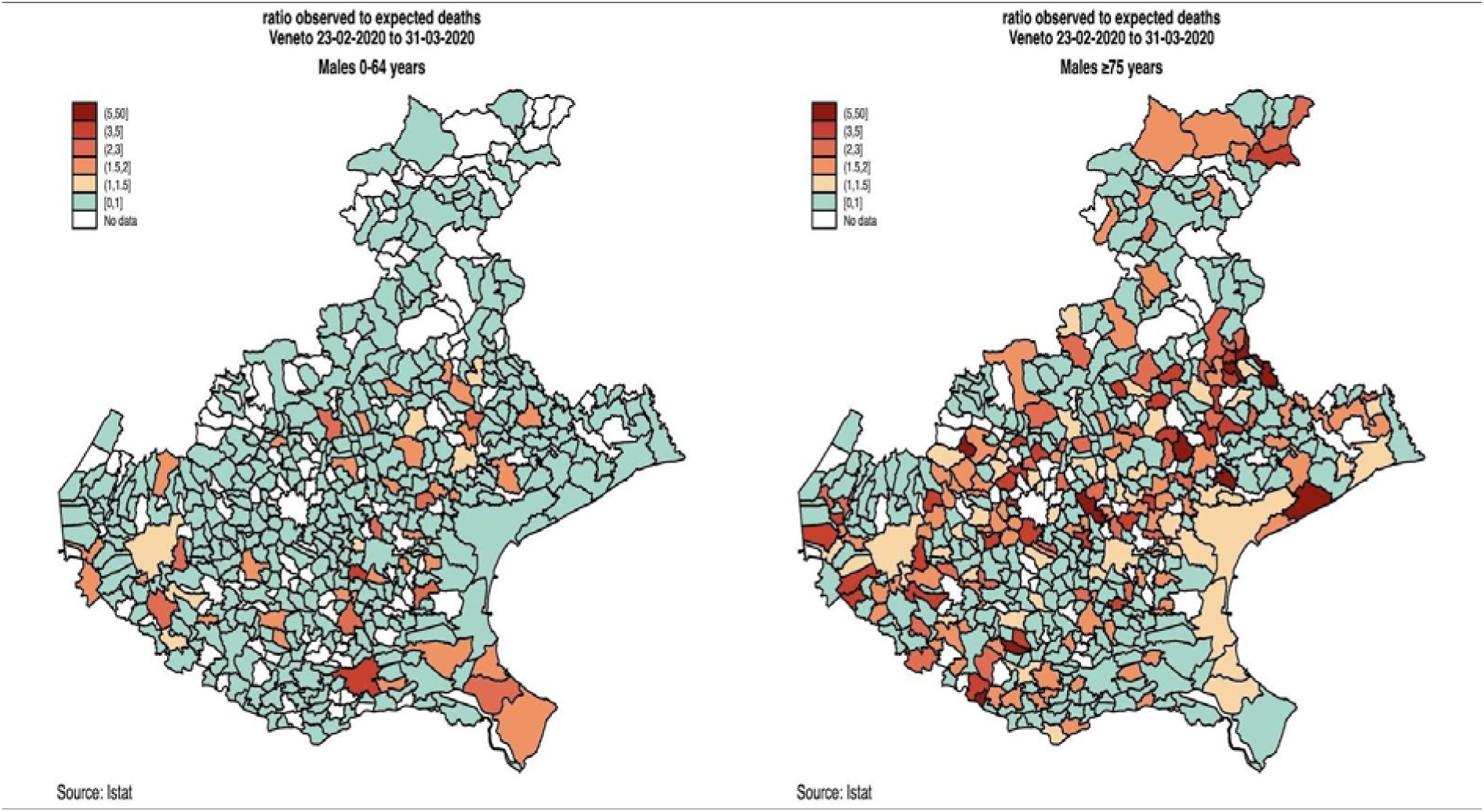
Relative mortality in males aged 0-64 (left) and ≥75 years (right) in Veneto region. Municipalities are depicted with different colors according to the magnitude of their excess mortality. Green refers to municipalities with a relative mortality ≤1, that is when observed deaths are lower than or equal to the expected deaths. When the relative mortality is >1, increasing saturation of red is used, to reflect increasing values of relative mortality. Municipalities depicted in white are those for which no data was released by ISTAT.

As for the Emilia-Romagna region (Figure 3), it 1 is evident for both age groups an EM north-south gradient from the area bordering the Lodi and Cremona provinces or Lombardy, following the trajectory of the main regional route that connects almost all the major regional centers. Moreover, one local hotspot of COVID-19 can be seen in the east of the region, spreading from the municipality of Medicina, and another local critical area might be noticed in the southernmost municipalities, bordering the Pesaro province of the Marche region, where another known hotspot was located. Similarly to Lombardy and Veneto, EM in the ≥75 age group was remarkably higher than in the 0-64 age group.

**Figure 3.**
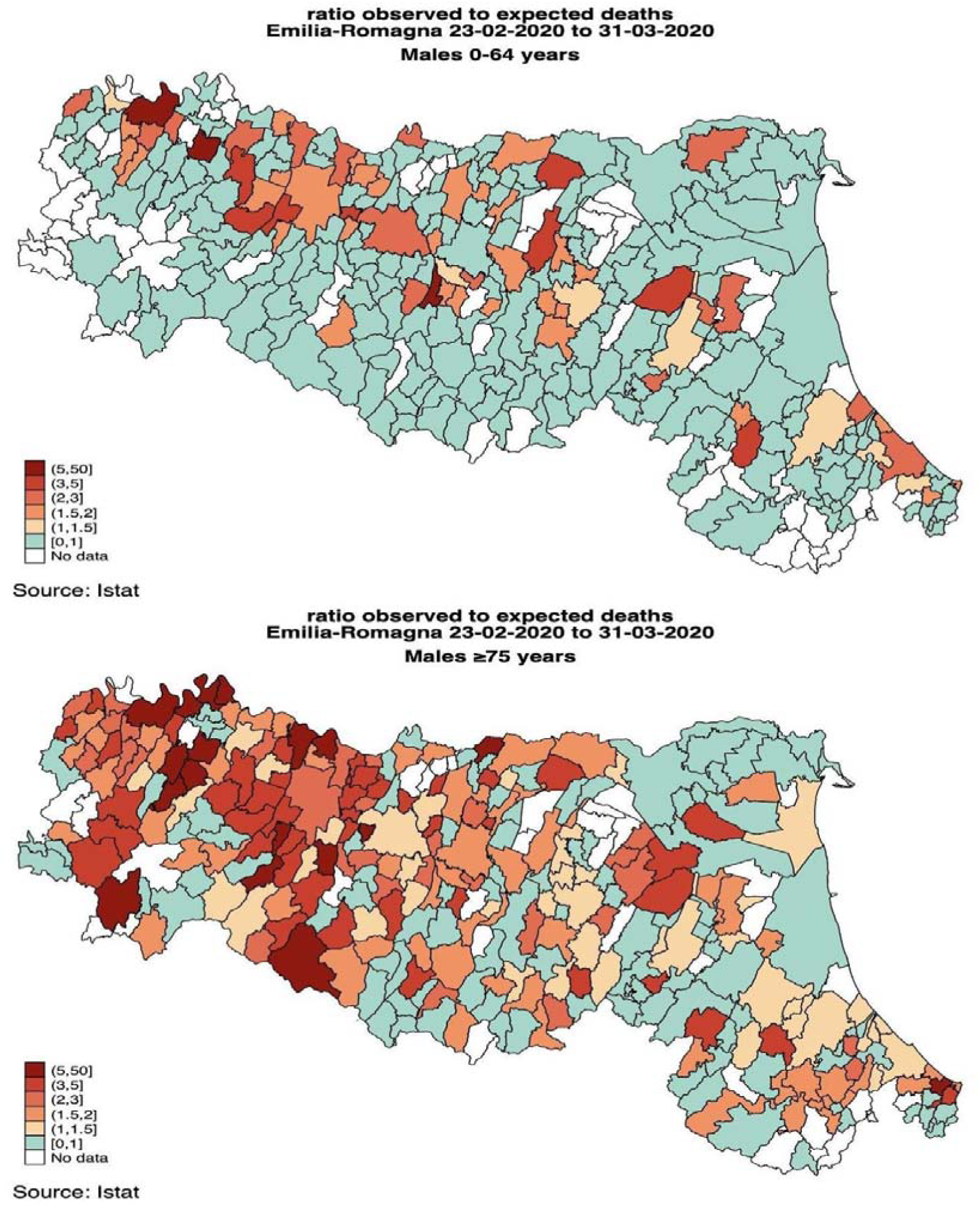
Relative mortality in males aged 0-64 (above) and ≥75 years (below) in Emilia-Romagna region. Municipalities are depicted with different colors according to the magnitude of their excess mortality. Green refers to municipalities with a relative mortality ≤1, that is when observed deaths are lower than or equal to the expected deaths. When the relative mortality is >1, increasing saturation of red is used, to reflect increasing values of relative mortality. Municipalities depicted in white are those for which no data was released by ISTAT.

Comparing the epidemic period with the first five weeks of the year, in the large majority of municipalities of Lombardy and Emilia-Romagna, and to a lesser extent in Veneto, mortality in 2020 before the COVID-19 outbreak was lower than expected (data not shown). This trend has been observed in many countries worldwide (EuroMOMO, 2020).

## 4. Discussion

In this study we found that excess mortality due to COVID-19 in Italy has an inhomogeneous spatial and demographic distribution in the most affected regions of Lombardy, Emilia-Romagna and Veneto, and within each region between the different municipalities.

Our study highlighted that COVID-19 related EM was similar between males and females for the ≥75 age group, while in the other age groups it was higher in males. Whether SARS-CoV-2 has a higher rate of severe clinical manifestations and mortality among older people will be clarified when more studies on the characteristics of patients will be available. However increasing age and pre-existing conditions such as diabetes, obesity and cardiovascular diseases could play an important role (Onder et al., 2020; Italian Superior Institute of Health, 2020).

The spread of COVID-19 epidemic clearly originated from specific areas and initially affected the surrounding municipalities. The epidemic was spread by individuals living in a community, and circulated across the regions through the main commercial and commuting routes, as suggested by other analyses (Adnkronos, 2020; Gloria, 2020; Golinelli et al., 2020; Sebastiani, 2020). Some areas distant from the principal epicenters most likely benefited from the lockdown measures enforced nationwide before the epidemic spread could massively reach them.

The different RHS organizations must be taken into account when analyzing the responses to the epidemic. Lombardy has a Regional Health System based on a strong hospital-centered approach, with many excellences in secondary and tertiary care. The Veneto region has a RHS with a solid primary care level; Emilia-Romagna is somewhere in between, but its health care system is more similar to the one in Veneto (Cicchetti et al., 2020). When SARS-CoV-2 started spreading in the town of Vo’, the RHS in Veneto and the policies adopted at the regional and local level allowed the region to quickly react. Adopting mainly a community based approach, a multitude of actions were taken at once: the municipality of Vo’ was immediately declared a red zone, massive testing (even if at the beginning of the outbreak test capacity is usually low (Ruan, 2020), isolation of cases, rapid contact tracing. In Vo’, all people, also the asymptomatic, were tested (Crisanti and Cassone, 2020; Lavezzo et al., 2020). Importantly, Veneto largely relied on home care assistance, limiting hospital admissions to the most severe cases and started early testing of the healthcare workers operating in the community and in the hospitals. In Emilia-Romagna, the community-based approach was used to contrast the epidemic as well, but Piacenza, where the first outbreak of the region was registered, originating from the nearby hotspot of Codogno, still remains one of the most affected municipalities in Italy. Probably the virus had already extensively spread when the first case was found, and the community services were not equipped to promptly react, being overwhelmed by the high volumes of contemporary cases. Also, it could be that the municipality was not quickly declared a red zone, when there was the suspicion that the number of cases still undetected might have been dangerously high. Nevertheless, the RHS was able to adapt and started being more effective in mitigating the spread of the virus, following the same approach used by Veneto, particularly leaning towards home care assistance and relying on general practitioners to implement an active surveillance system among their patients with phone calls to monitor their symptoms.

On the contrary, the organizational structure of the RHS in Lombardy that promotes a hospital centered approach at the expenses of the community-based services, may have contributed to exacerbate the criticalities presented by COVID-19. Thereto, the region did not address the outbreak adopting the same plan in municipality experiencing similar circumstances: while the towns around Lodi were quickly declared red zones when the first cases were identified, thus containing the spread of the virus, the outbreak of the virus in a hospital in the province of Bergamo, didn’t lead to the establishment of a confinement, that was implemented only when the national lockdown was enforced (De Luca, 2020). This contributed to make the Bergamo area one of the most hit areas in Europe. Especially at the beginning of the epidemic, most people were admitted to hospitals. The dramatic flow of patients saturated the intensive care units, forcing doctors to decide who could go forward (Rosenbaum, 2020). In the hospital setting, the virus was spread not only by patients, but also by healthcare workers, who could not always rely on appropriate personal protective equipment (PPE) risking their lives while doing their job.

Analyzing the deaths’ toll in Lombardy, Veneto and Emilia-Romagna, we argue that the best way to address epidemics is through proactive medicine at a community-based level, opposite to reactive medicine at a hospital level. Moreover, to foster discussions and more in depth analysis on the EM due to COVID-19 infections, we hypothesized several factors that could have equally affected the infections and mortality in the three regions: household transmission, which led to the most vulnerable being exposed when a family member got the infection; congestion of the healthcare system; late diagnosis of the healthcare workers for COVID-19; limited number of executed swabs; inadequate contact tracing; public transportation; inadequate community based interventions.

At the regional level, the difference in adopted strategies certainly reflects different RHS’s organization and operational approaches. In tackling the epidemic, the Veneto and Lombardy regions adopted two opposite strategies regarding, for example, the recommendations on testing. As of March 31, Veneto performed 21.6 swabs per 1,000 population (4,905,854 inhabitants) and, whenever possible, tested also asymptomatic people; Lombardy (10,060,574 inhabitants) performed 11.3 swabs per 1,000 population, testing mainly patients with more severe clinical symptoms, as instructed by the Italian Ministry of Health in that moment. On this topic, Emilia Romagna had a more similar approach to Lombardy, performing 12.2 swabs per 1,000 population on a total population of 4,459,477 inhabitants (Italian Civil Protection Department, 2020; Italian Institute for National Statistics (ISTAT), 2020a; Onder et al., 2020).

At the local level, the reason of higher COVID-19 mortality in some specific municipalities can be debated, as several different causes other than the distance from the hotspots or a possible bias in estimation or some casual local factors might be involved. Possible explanations for a marked difference in excess mortality between close municipalities could be the presence of gathering places (i.e. sport venues, cinemas), or of some industrial/commercial activities, care and nursing homes or of particular events that may have taken place before the lockdown. In the municipality of Medicina (Emilia Romagna), a gathering at a bowls club was the cause of the rapid spread of the virus; almost all the people present got infected, and all but one died. Medicina immediately became a red zone (Regione Emilia-Romagna, 2020), successfully narrowing the afflicted area. This demonstrates that it is important to contrast the spread of the virus at the smallest possible level and in the most accurate possible way.

Several municipalities showed a lower increase of COVID-19 related deaths or even a reduced mortality. This can be likely attributed to a smaller incidence of deaths for causes such as accidents or occupational injuries, favored by the lockdown. On the other hand, the high mortality figures especially for people aged ≥75 may also have been favored by the lower incidence of influenza observed this year in Italy, compared to last year, which left more elderly people exposed to COVID-19 (Bella, 2020).

Our study suggests that the difference in the response to the epidemic can be the result of policies adopted both at the regional and at the municipality level. Most of the studies conducted on this epidemic (Bucci et al., 2020; Buonanno et al., 2020; Modi et al., 2020; ScienzaInRete, 2020) have tried to explain its course considering the regional policies, as so did we, but, as an added value, in the present study we explored also the pattern of the epidemic within regions and noticed that, net of regional policies, even nearby municipalities showed different excess mortality rates. Accordingly, the causes of such differences should be sought not only at the national and the regional policies, but also in local factors which cannot be brought to light if data are analyzed as regional or higher aggregates. Therefore, focusing on the regional and municipality level could be more helpful than a regional level analysis alone and we advocate that our approach may help to sned some light un me areas where the death xuii was highest arid mus trigger Tuimei investigations on some speclTlc/lucal dynamics uT the outbreak.

Our study presents some caveats. First, it is based on incomplete territorial coverage, therefore our conclusions could have been partially different if data for all municipalities was available. However, our data cover >87% of Italian municipalities, which is a relevant coverage. We used linear regression to derive a mortality trend from 5 years, which could not always be adequate as it may be leveraged by anomalous mortality figures in the first or in the last year of the time interval used for prediction. By using linear regression, we intended to capture a trend in mortality where it exists, and in these cases its prediction is more accurate than the projection of the average mortality observed in the previous five years. In addition, we reduced potential confounding by estimating excess mortality within subgroups defined by age class and gender. Nevertheless, our results should be considered as provisional estimates until official data of verified quality covering all municipalities will be released and made available to researchers for further investigations.

## 5. Conclusions

Our approach could be used to generate or to confirm hypotheses regarding the flow of epidemics starting from the main originating sites, and to detect affected loci that are distant from these initial affected areas. Analyses are fundamental at the regional and subregional level. Identifying the municipalities where the mortality burden was higher, and the pathways used by the virus to spread may help to concentrate efforts in understanding the reasons why this happened and to identify the frailest areas in case of the occurrence of a second epidemic outbreak. Whether higher excess mortality depended on lack of preparedness, or on a particularly old age structure, or on inefficacy in the enforcement of containment measures, this should lead to different types of interventions in the preparation of a possible second wave of COVID-19.

The phase of coexistence with the virus will be as challenging as the phase of the outbreak. An adequate organization of the health services is imperative.

Our study demonstrates that nearby municipalities within each region may show highly different mortality levels, despite being under common regional health policies, therefore further studies are necessary to analyze more in depth the local determinants of COVID-19 spread.

## Data Availability

All the data referred to are public data given by the Italian National Institute of Statistics.
Supplementary data will be made available at request

## DECLARATIONS

### Availability of data and material

Data are publicly available.

### Ethics approval

Not applicable.

